# Associations of autistic traits, sleep/circadian factors, and mental health

**DOI:** 10.1101/2025.06.12.25329485

**Authors:** Neha Deshpande, Siddhi Nair, Elise Taylor, Eus Van Someren, Sarah Laxhmi Chellappa

**Affiliations:** Centre for Innovation in Mental Health, School of Psychology, Faculty of Environmental and Life Sciences, University of Southampton, Southampton, United Kingdom; Netherlands Institute for Neuroscience, Department of Sleep and Cognition, Amsterdam, the Netherlands; Departments of Integrative Neurophysiology and Psychiatry, Center for Neurogenomics and Cognitive Research, VU University, Amsterdam UMC, Amsterdam Neuroscience, Amsterdam, the Netherlands

**Keywords:** Autism, sleep complaints, insomnia, sleep quality, chronotype, social jetlag, mental health, depression, quality of life

## Abstract

**Background:** Autistic individuals experience a heightened risk of depression and lower quality of life; however, it remains to be established whether disrupted sleep and circadian factors mediate this increased risk.

**Objectives:** We assessed whether disruption of self-reported sleep and circadian factors mediate the associations of autistic traits with depression symptom severity and quality of life.

**Methods:** 838 participants (mean: 52.8 [*SD* = 1.3] years, 70% females) from a large-scale observational survey (Netherlands Sleep Registry) completed the Autism Quotient Scale (AQ), Hospital Anxiety and Depression Scale Cantril Ladder quality of life, Insomnia Severity Index, Pittsburgh Sleep Quality Index, and the Munich Chronotype Questionnaire.

**Results:** \Higher autistic traits were associated with a trend for higher depression symptom severity (*p* = 0.06), significantly lower quality of life (*p* < 0.001), higher insomnia severity (*p* < 0.001), lower sleep quality (*p* < 0.001), a trend for late chronotype (*r* = 0.04, *p* = 0.06), but not social jetlag (*r* = 0.02, *p* = 0.21). Insomnia severity and chronotype partly mediated the association of autistic traits and depression symptom severity (standardized beta = −0.02, 95% CI = [−0.04, 0.00]), and the association of autistic traits and quality of life (standardized beta = −0.02, 95% CI = [−0.04, 0.00]).

**Conclusion:** Autistic traits were associated with depression severity and lower quality of life, mediated by insomnia symptom severity and chronotype. Future studies targeting insomnia complaints and late chronotype in this population may help alleviate their mental health complaints and increase quality of life.

## INTRODUCTION

Autism often co-occurs with mental disorders, particularly depression, such that up to 40% of autistic individuals report at least one episode of depression throughout their lifespan compared to neurotypically developing individuals (Hollocks et al. 2019). Moreover, autistic individuals often report lower quality of life (e.g., life satisfaction and psychological wellbeing), compared to neurotypically developing individuals (Ayres et al. 2018, Park et al. 2019). Despite this major public health concern, it remains to be established what biological factors contribute to these associations. Importantly, most studies focus on children and adolescents, with little known about whether autistic middle-aged and older adults share a similar risk of depression and low quality of life.

Sleep disturbances are common among autistic children and persist throughout their development and into adulthood compared to neurotypically developing adults (Humphreys et al. 2014). Meta-analysis of 8 datasets (194 autistic adults and 277 neurotypically developing adults) indicates that autistic adults had significantly higher levels of impairment for self-reported sleep quality, including lower sleep efficiency (SMD = −0.87, CI = −1.14 - 0.60) (Morgan et al. 2020). Similar findings were described for objective sleep, including longer sleep onset latency (PSG-assessed, SMD = 0.86, CI = 0.29-1.07) and wake after sleep onset (wristworn wearable-assessed) (SMD = 0.57, CI = 0.28-0.87) (Morgan et al. 2020). Likewise, up to 25% of autistic children and adolescents report insomnia symptoms compared to neurotypically developing young people (Diaz-Roman et al. 2018). Autistic children and adolescents may also experience circadian disturbances, including late chronotype and more social jetlag (Taylor et al. 2024), with similar findings for chronotype in autistic adults (Harris et al. 2023). Sleep and circadian rhythm disruption (SCRD) are bidirectionally associated with a plethora of mental disorders (Meyer et al. 2024). However, as most studies primarily focus on neurotypically developing individuals, it remains unclear if SCRD mediates the increased risk of depression and lower quality of life in individuals with autistic traits.

This study aimed to establish whether disruption of self-reported sleep (e.g., insomnia symptom severity and self-reported sleep quality) and circadian factors (e.g., chronotype and social jetlag) mediate the association of autistic traits with depression severity and low quality of life in a large-scale cross-sectional study.

## METHODS

### Participants

Data included here are from the Netherlands Sleep Registry (NSR), an online survey platform dataset with > 10,000 individuals (aged 18 and older) from the general population (www.slaapregister.nl). Participants were recruited via media, advertisements, and flyers distributed at healthcare institutions and conventions. Participation was voluntary, and all data were recorded anonymously (Blanken et al. 2019). The only inclusion criterion was age> 18 years. Participants completed a variable number of selected questionnaires at their convenience, resulting in a varying number and set of completed questionnaires across participants. Here, 838 participants fully completed questionnaires about autism, insomnia, sleep quality, chronotype, social jetlag, depression, and quality of life.

### Ethics approval

Volunteers of the NSR were not exposed to interventions or behavioral constraints, and, therefore did not to fall under the Dutch Medical Research Involving Human Subjects Act, as judged by the Medical Ethical Committee of the Academic Medical Center of the University of Amsterdam as well as the Central Committee on Research Involving Human Subjects (CCMO), The Hague, The Netherlands. Participants digitally signed informed consent with anonymized use of their data. Before performing this data analysis, we obtained ethical approval from the University of Southampton Research Ethics Committee (ERGO: 93647).

### Self-reported measurements

#### Autistic traits

The Autism Spectrum Quotient (AQ) is a self-administered instrument to assess adults with autistic traits according to the DSM-5 (Baron-Cohen et al. 2001). The AQ comprises 50 items, including social skills, attention switching, communication, imagination, and attention to detail. Participants rated these items on a 5-point ordinal Likert scale, ranging from not at all to very much. Higher AQ scores indicate higher autistic traits.

#### Depression and Quality of life

Hospital Anxiety and Depression Scale is a self-report questionnaire with seven items measuring depression (Zigmond et al. 1983). Each item is scored on a 4-point Likert scale with the scoring range varying from 0 to 21. Scores between 0-7 indicate normal levels, 8-10 are near-threshold abnormal, and 11-21 indicate depression severity. The Cantril’s Self-Anchoring Ladder of Life satisfaction measures life satisfaction by visualizing an adult’s life in the best possible manner and explaining dreams and hopes for the future. A score of four or below indicates ‘suffering’, and scoring more than seven equates to ‘thriving’. Higher scores indicate better life satisfaction and wellbeing.

#### Insomnia and Sleep quality

The Insomnia Severity Index (ISI) is a 7-item questionnaire on a 5-point Likert scale ranging from 0 (not at all) to 4 (very severe/very much). The total score ranges between 0-28 with the cutoff score for likely insomnia of least 10. The Pittsburg Sleep Quality Index (PSQI) assessed difficulties initiating and maintaining sleep. This 19-item self-report questionnaire includes sleep latency, sleep duration, subjective sleep quality, habitual sleep efficiency, sleep disturbances, daytime dysfunction, and use of sleeping medication. Scores range from 0 (no difficulty) to 3 (severe), with higher scores indicating low sleep quality.

#### Chronotype and Social jetlag

The Munich Chronotype Questionnaire (MCTQ) is a self-rated scale that assesses an individual’s entrainment of working and work-free days. It assesses chronotype by evaluating the corrected midsleep on free days, i.e., midpoint between sleep onset and offset on free days. Social jetlag was calculated using the difference between the midpoint of sleep on workdays and the midpoint of sleep on free days.

#### Data analysis

Data was analyzed using SAS 9.4. To standardize the dataset, all of the data were z-transformed. We applied Spearman correlations to assess the associations of autistic traits, depression symptom severity, quality of life, insomnia severity, self-reported sleep quality, chronotype, and social jetlag. Hierarchical regression models were performed to determine the effect of autistic traits on depression symptom severity and quality of life, while accounting for sociodemographic factors, insomnia severity, self-reported sleep quality, chronotype, and social jetlag. The first model accounted for sociodemographic variables (age, sex, marital status, educational level, employment) and the second model included all sociodemographic variables, as well as insomnia severity, sleep quality, chronotype, and social jetlag. Effect sizes were determined by the R^2^ values with 0.02, 0.13, and 0.26 indicating small, medium, and large effect sizes, respectively. We assessed potential multicollinearity by using methods such as correlation coefficients or Variance Inflation Factor. Finally, mediation analyses were conducted to identify whether sleep/circadian factors (i.e., sleep quality, insomnia severity, chronotype, and social jetlag) mediated depression symptom severity and quality of life in individuals with autistic traits. The first mediation model included the sleep/circadian predictors self-reported sleep quality, chronotype, social jetlag, and insomnia severity. The second model included the sleep/circadian factors and sociodemographic covariates. In both models, we included multiple possible mediators simultaneously. We present the standardized beta values based on the types of variables, including exposures, mediators, and outcomes (Abrar et al. 2024).

## RESULTS

### Correlational analysis

Higher autistic traits were associated with a trend for higher depression symptom severity (*r* = 0.04, *p* = 0.06), and significantly lower quality of life (*r* = −0.20, *p* < 0.001). Similarly, higher autistic traits were significantly associated with higher insomnia severity (*r* = 0.08, *p* < 0.001), and lower sleep quality (*r* = 0.08, *p* < 0.001). Additionally, we observed a trend between autistic traits and late chronotype (*r* = 0.04, *p* = 0.06), but not for social jetlag (*r* = 0.02, *p* = 0.21).

### Hierarchical regression models

For the analyses of depression, none of the sociodemographic predictors significantly contributed to depression severity (model 1: R^2^ = 0.02; all p > 0.1; model 2: R^2^ = 0.07; all p > 0.1). However, self-reported sleep quality (model 2: R^2^ = 0.07; p = 0.01) and autistic traits (model 2: R^2^ = 0.07; p = 0.02) significantly contributed to depression severity (**Table 1**). The combined effects of sleep/circadian predictors, sociodemographic factors, and autistic traits had a small effect (7% of the variance; model 2) (**Table 2**).

**Table 1.**
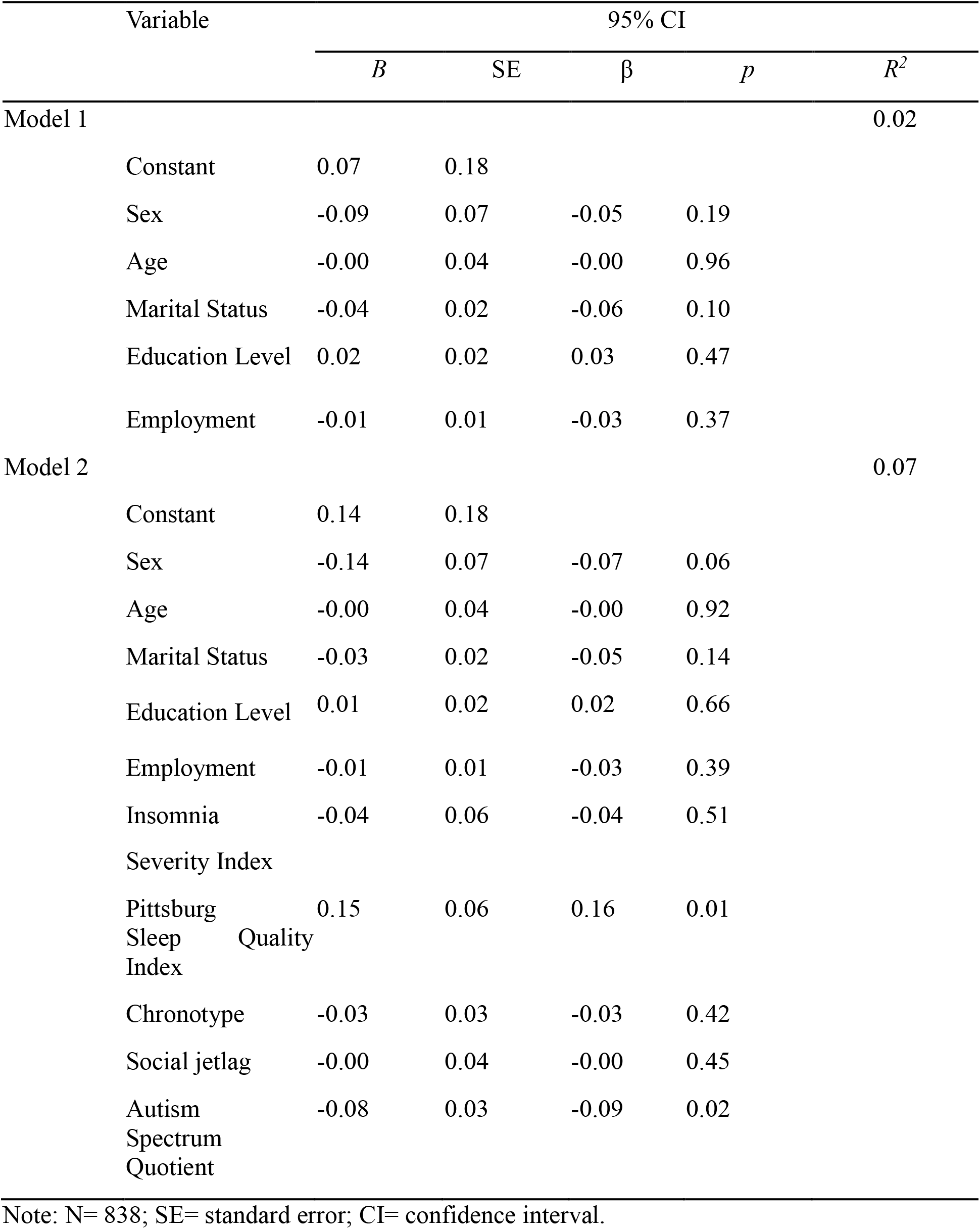
Hierarchical Regression Results for depression symptom severity.

**Table 2.** Hierarchical Regression Results for quality of life (Cantril Ladder).

| Variable | 95% CI | | | | $R^2$ |
| --- | --- | --- | --- | --- | --- |
| | $B$ | SE | $\beta$ | $p$ | |
| Model 1 |  |  |  |  | 0.20 |
| Constant | 1.47 | 0.08 |  |  |  |
| Sex | -0.09 | 0.03 | -0.10 | 0.004 |  |
| Age | -0.04 | 0.02 | -0.09 | 0.006 |  |
| Marital Status | 0.03 | 0.01 | 0.09 | 0.003 |  |
| Education Level | -0.03 | 0.01 | -0.09 | 0.008 |  |
| Employment | 0.01 | 0.01 | 0.04 | 0.28 |  |
| Model 2 |  |  |  |  | 0.25 |
| Constant | 1.44 | 0.08 |  |  |  |
| Sex | -0.07 | 0.03 | -0.07 | 0.03 |  |
| Age | -0.04 | 0.02 | -0.09 | 0.007 |  |
| Marital Status | 0.03 | 0.01 | 0.09 | 0.006 |  |
| Education Level | -0.02 | 0.01 | -0.08 | 0.018 |  |
| Employment | 0.01 | 0.01 | 0.04 | 0.28 |  |
| Insomnia | 0.09 | 0.03 | 0.20 | 0.001 |  |
| Severity Index |  |  |  |  |  |
| Pittsburg Sleep Quality Index | 0.00 | 0.03 | 0.01 | 0.89 |  |
| Chronotype | 0.01 | 0.01 | 0.02 | 0.52 |  |
| Social jetlag | -0.00 | 0.04 | -0.00 | 0.47 |  |
| Autism Spectrum Quotient | 0.03 | 0.01 | 0.08 | 0.03 |  |

Note: N= 838; SE= standard error; CI= confidence interval.

For the analyses of quality of life, the sociodemographic predictors sex, age, marital status, and educational level significantly contributed to quality of life (model 1: R^2^ = 0.20; all p < 0.02). Similarly, insomnia severity and autistic traits significantly contributed to quality of life (model 2: R^2^ = 0.25; respectively, p = 0.001, p = 0.04, and p = 0.03). The combined effects of sleep/circadian predictors, sociodemographic factors, and autistic traits had a moderate effect (25% of the variance; model 2) (**Table 2**).

### Mediational analysis

For the analyses of depression, the association of autistic traits with depression severity was partly mediated by insomnia severity and chronotype before and after including sociodemographic factors (standardized beta = −0.02, bootstrapped 95% CI = [−0.04, - 0.00]). Similarly, for the analyses of quality of life, the association between autistic traits and quality of life was partly mediated by insomnia severity and chronotype before and after including sociodemographic factors (standardized beta = −0.02, bootstrapped 95% CI = [−0.04, 0.00]).

## DISCUSSION

We showed that autistic traits were associated with higher depression symptom severity, lower quality of life, insomnia severity, and lower self-reported sleep quality. Additionally, insomnia severity and late chronotype partly mediated the association of autistic traits with depression symptom severity and with quality of life.

A meta-analysis of 35 studies, including 29 studies measuring depression (n = 26,117) indicated that the pooled estimation of current and lifetime prevalence for major depressive disorder in young autistic adults were 23% and 37%, respectively, nearly twofold higher than the risk observed in the general population (Hollocks et al. 2019). Moreover, a meta-analysis with 10 studies (n= 486 autistic middle-age and older adults and 17,776 controls) indicated lower quality of life (including physical health, psychological health, social relationships, and environment wellbeing) in autistic adults, with a large effect (Cohen’s d = −0.96) (van Heijst et al. 2015). Such an increased risk of depression and lower quality of life in adulthood may help explain why autistic individuals are at a heightened risk of increased loneliness, employment difficulties (Hedley et al. 2017), and suicidal ideation and attempts (Cassidy et al. 2018). Our findings show similar associations with depression symptom severity and lower quality of life in individuals with autistic traits from the general population.

Importantly, we show that these associations are mediated by disruption of sleep and circadian factors, particularly insomnia symptom severity and late chronotype. Insomnia and low sleep quality in autism are significantly biological in origin, as evidenced by a robust genetic correlation of 0.62 between autism (n=15,279 child and adolescent twin pairs) and difficulties initiating and maintaining sleep (Taylor et al. 2022). In a similar vein, a genome-wide association study showed that insomnia risk genes are enriched in autistic children and adolescents compared with neurotypically developing people (Tesfaye et al. 2022). Up to 30% of autistic individuals may exhibit late chronotype and/or late dim light melatonin onset (Carmassi et al. 2019), attenuated amplitude of endogenous circadian melatonin levels, and irregular melatonin release patterns (Bouteldja et al. 2024). Genetic studies have shown polymorphisms in circadian CLOCK genes and genes involved in melatonin synthesis in autism, which have been linked to increased insomnia symptoms in autistic children and adolescents as compared to those with neurotypical development (Pagan et al. 2014). Given the bidirectional relationship of SCRD, depression and wellbeing, such disruptions may likely contribute to the higher risk of mental health problems experienced by autistic people.

Limitations of this study include the age range (middle-aged adults) and (mostly) white females, which limits the generalizability of the findings. We did not assess objective markers of sleep quality (e.g., wristworn-wearables for estimating sleep-related parameters) and circadian phase (e.g., dim light melatonin onset). Lastly, the cross-sectional design of this study does not infer causality; therefore, longitudinal studies are required to establish a causal role of e.g., insomnia in the association of autism and mental health problems.

In conclusion, our study shows that autistic traits were associated with higher depression symptom severity and lower quality of life, and this effect was mediated partly by insomnia symptom severity and late chronotype. Future studies targeting insomnia complaints and later circadian timing in autistic individuals may help mitigate mental health complaints, with ramifications for their quality of life.

## Acknowledgements

The Netherlands Sleep Registry study was co-funded by the European Union (ERC-AdG Overnight, 101055383). Views and opinions expressed are however those of the authors only and do not necessarily reflect those of the European Union or the European Research Council. Neither the European Union nor the granting authority can be held responsible for them. The funder had no role in the design of the study; in the collection, analysis, or interpretation of data; in the writing of the manuscript, or in the decision to publish it.

## Author Contributions Statement

Conceptualization: E.V.S. Funding acquisition: E.V.S. Investigation: E.V.S. and S.L.C. Visualization: S.N., N.D., E.V.S., and S.L.C. Project administration: S.N., N.D., and S.L.C. Supervision: S.L.C. Data curation: S.N., N.D., E.V.S., and S.L.C. Formal analysis: S.N., N.D., and S.L.C. Validation: E.V.S., and S.L.C. Writing—review and editing: S.N., N.D., E.T., E.V.S., and S.L.C.

## Data Availability Statement

The data that support the findings of this study are available from the shared senior author and Principal Investigator, [E.V.S.], upon reasonable request.

